# Synchronous Group-Based Tele-Exercise versus Community-Based Interventions: Effects on Physical Functioning and Adherence in Older Adults at Risk of Falls - A Randomized Controlled Trial

**DOI:** 10.1101/2024.06.16.24308943

**Authors:** Karly O. W. Chan, Peter P. Yuen, Ben Y.F. Fong, Vincent T.S. Law, Fowie S.F. Ng, Janet L.C. Lee, I.S. Cheung, Tommy K.C. Ng, Wilson C.P. Fung

## Abstract

**Background:** Falls are common in older adults and can cause serious harm. Tele-exercise offers a promising alternative to traditional programmes, especially for those with mobility or access limitations. However, existing studies are limited by high-risk-of-bias, non-representative samples and inadequate sample size estimation, necessitating rigorous research. This study addressed these gaps by evaluating the effectiveness of synchronous group-based tele-exercise (TE) versus community-based (CB) exercise in improving physical function, exercise adherence and maintenance among older adults at risk of falls.

**Methods:** Ninety-four community-dwelling older adults from 10 Hong Kong community centres were randomized to TE or CB groups. Both received modified Otago exercise training for 3 months with 9-month follow-up. Fall and functional outcomes included Fall Efficacy Scale-International (FES-I), 6-meter walk test, Timed Up and Go (TUG), Berg Balance Scale (BBS), Functional Reach Test (FRT), and Appendicular Skeletal Muscle Mass Index (ASMI), assessed at baseline, 3, 6, and 12 months. Exercise experience and maintenance outcomes included exercise adherence and Physical Activity Scale for the Elderly (PASE-C) and physical activity level. Analysis used a modified intention-to-treat approach.

**Results:** Between group analysis revealed that TE had significantly higher PASE-C scores at 12 months (p<0.05, ES=0.37) and greater light activity minutes at 6 months (p<0.05, ES=0.33). In contrast, CB demonstrated significantly higher adherence (90% vs. 80%, p=0.01) and lower dropout rate (0% vs. 10.4%, p<0.01). No significant between-group differences were observed in fall risk or physical function outcomes. Within group analysis revealed that both groups improved in fall risk and physical function to varying extents. No serious adverse events occurred.

**Conclusions:** Synchronous group-based tele-exercise is as effective as face-to-face community training for improving fall risk and physical function among older adults at risk of falls. Notably, tele-exercise demonstrated a slight advantage in sustaining long-term exercise maintenance. This may be attributed to participants becoming accustomed to exercising at home during the initial three-month intervention, making it easier to continue training independently over the following nine months. However, the higher dropout rate in the tele-exercise group raises concerns. These findings support tele-exercise as a viable and accessible alternative for promoting long-term physical activity in older adults.

**Trial registration number:** ChiCTR2200063370

## Background

Falls among older people are a significant concern, with a global prevalence of 26.5% [1]. Falls can lead to substantial costs, ranging from 2,044 to 25,955 USD per fall victim, 1,059 to 10,913 USD per fall event, and 5,654 to 42,840 USD per fall-related hospitalization [2]. Hospitalizations due to falls are five times more common than those resulting from other injuries [3]. Falls also have adverse consequences such as hip fractures, rehospitalization, extended hospital stays, and increased mortality rates [4, 5]. Therefore, implementing preventive strategies is vital in mitigating age-related impairments and promoting independence in older adults.

Traditional exercise interventions have shown promise in reducing fall risk and preventing fall-related injuries [6]. However, older adults face significant challenges due to limited exercise space in the community and travel constraints [7], especially for those who are susceptible to falls. To address these challenges, tele-exercise has emerged as a viable alternative. Tele-exercise encompasses various forms, including video, audio, or text communication technologies, delivered synchronously or asynchronously [8, 9]. Chan et al. [10] reported promising results in reducing falls among community-dwelling older adults through telehealth combined with exercise interventions and highlighted the safety, feasibility, and acceptability of telehealth interventions. The presence of real-time guidance and supervision of video conferencing-based online exercise programmes was found to be highly beneficial for older adults [7]. Granet et al. [11] demonstrated that a higher ratio of live training resulted in greater improvements in muscle function and a lower drop-out rate compared to a higher ratio of recorded sessions, emphasizing the relevance of synchronous delivery of tele-exercise.

The existing studies on synchronous tele-exercise for community dwelling older adults and older adults at risk of falls have limitations. They predominantly focused on older adults who already had an internet connection and the necessary devices for online exercise programmes as inclusion criteria [11–13]. Granet et al. [11] noted that the study samples were highly educated, with over 67% of participants having attained a university level of education, which may limit the generalizability of the results to the broader population. In addition, the lack of proper sample size estimation in some studies also highlights the need for improved study quality [11, 13].

In a recent systematic review conducted by Gamble et al. [8], the methodological quality of the evidence from 26 randomized controlled trials about the effectiveness of telerehabilitation such as tele-exercise for community dwelling older adults was critiqued. Among the included trials, four specifically targeted older adults with an increased risk of falls [13–16]. It is noteworthy that three of these studies[13, 15, 16] exhibited a high risk of bias based on the revised Cochrane Risk-of-Bias 2 tool [17], emphasizing the necessity for more rigorous and controlled investigations in this particular field. Interestingly, only two of these studies incorporated a videoconferencing setup [13, 15], which is an essential component of synchronous tele-exercise.

Therefore, the aim of the current study was to address these methodological limitations by investigating the effectiveness of synchronous tele-exercise on physical functioning, exercise adherence and exercise maintenance among community-dwelling older adults at risk of falls in comparison with a community-based group.

## Methods

### Study design

This was a 3-month single-blind, randomized controlled trial (RCT) comparing synchronous group-based tele-exercise training (TE) with face-to-face community-based exercise training (CB) for community-dwelling older people at risk of falls. Both groups of participants were followed for the subsequent 9 months. More detailed information, including details of the outcome measures and intrinsic differences between the two groups, can be obtained from our study protocol [18]. Reporting follows the CONSORT 2025 statement [19].

### Research ethics

Approval for the study was granted by the Ethics Committee of the College of Professional and Continuing Education (RC/ETH/H/0029). The study was registered in the Chinese Clinical Trial Registry (ChiCTR2200063370) on 5 September 2022. In accordance with the WMA declaration of Helsinki, informed consent was obtained from all participants.

### Sample size calculation

A review of digital technology-based OEPs reported between-group effect sizes ranging from 0.43 to 0.73 for balance, muscle strength, and fall efficacy in programmes lasting more than 12 weeks [20]. As the CB group is a positive control, the expected between-group effect size for the TE versus CB is likely smaller than that observed in studies with inactive or placebo controls. To avoid underpowering the study, we selected a conservative effect size of 0.367, based on a preliminary study by Benavent-Caballer et al. [21] which evaluated the Time Up and Go (TUG) Test as the primary outcome.

Sample size calculations were conducted using G*Power software, version 3.1.9.7 (Heinrich-Heine-Universität Düsseldorf, Düsseldorf, Germany), With a confidence level of 95%, a power of 80%, and an estimated attrition rate of 20% (including mortality) [22], a minimum sample size of 92 participants was determined to be necessary.

### Participants and eligibility criteria

The participants were recruited from 10 elderly community centres located in various locations across Hong Kong. The inclusion and exclusion criteria are listed in table 1.

**Table 1.**
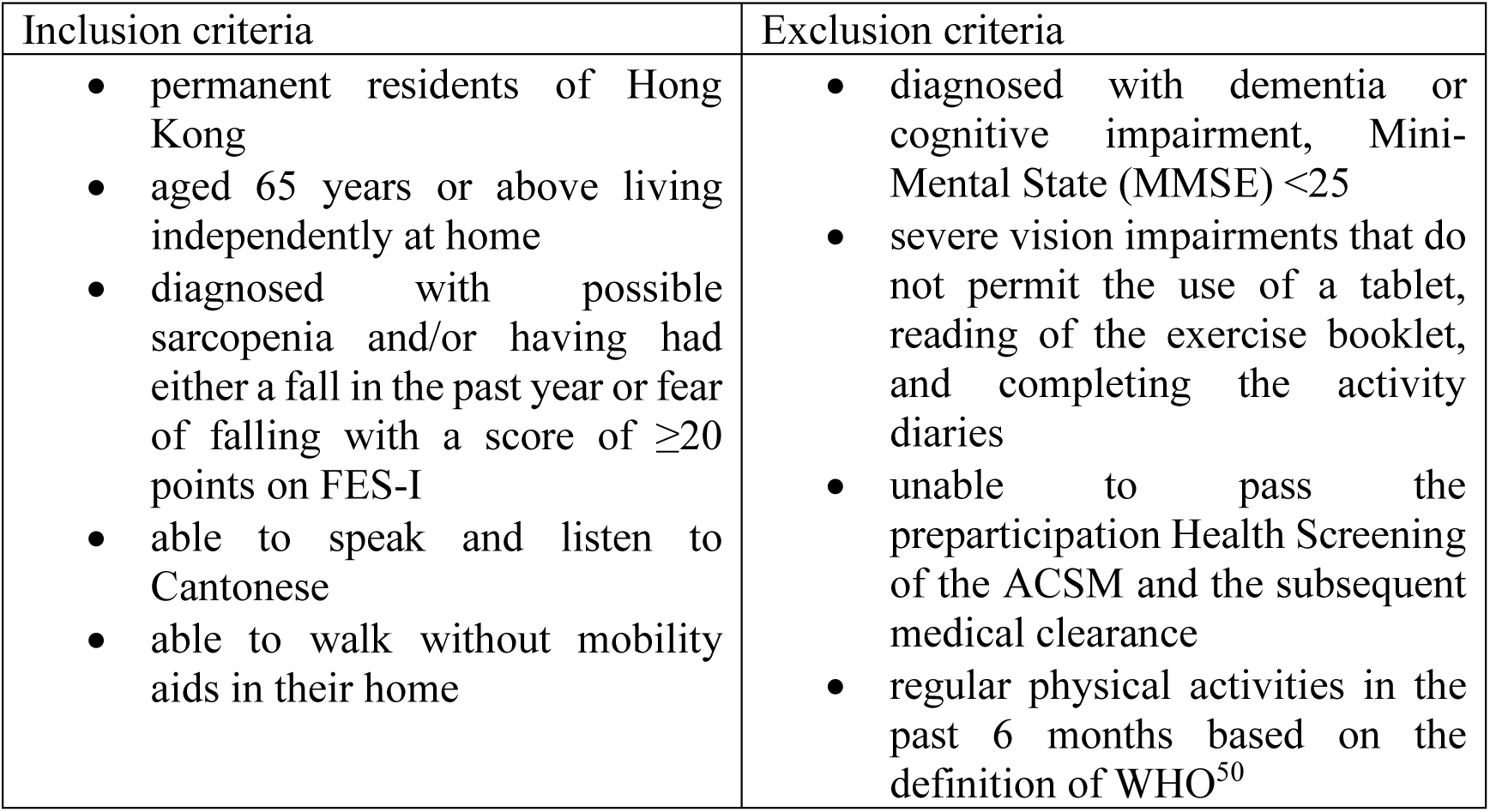
Inclusion and exclusion criteria (adopted from Chan et al. [18]).

| Inclusion criteria | Exclusion criteria |
| --- | --- |
| • permanent residents of Hong Kong<br>• aged 65 years or above living independently at home<br>• diagnosed with possible sarcopenia and/or having had either a fall in the past year or fear of falling with a score of $\geq 20$ points on FES-I<br>• able to speak and listen to Cantonese<br>• able to walk without mobility aids in their home | • diagnosed with dementia or cognitive impairment, Mini-Mental State (MMSE) $< 25$<br>• severe vision impairments that do not permit the use of a tablet, reading of the exercise booklet, and completing the activity diaries<br>• unable to pass the preparticipation Health Screening of the ACSM and the subsequent medical clearance<br>• regular physical activities in the past 6 months based on the definition of WHO<sup>50</sup> |

### Randomization and blinding

Ten community centres were coded and then randomly assigned to either the TE or CB with a 1:1 allocation in a concealed, blinded fashion to minimize cross-over contamination between the groups. Randomization procedure and allocation sequence were supervised by an independent assistant. The project assistant enrolled participants in the community centres. Trained outcome assessors, who were blinded to group assignments, performed the assessments. Participants were aware of their unique training modes but had no contact with participants from other groups due to different locations. Instructors and researchers were not blinded and were instructed not to discuss group assignments with the participants and outcome assessors.

### Intervention programmes

The intervention programme have been described in detail in the published protocol [18]. All participants engaged in exercise training sessions three times a week for a duration of three months, either through tele-exercise or face-to-face at the community centres. Each session lasted for 45 minutes. The participants in both groups were divided into classes consisting of 8 to 11 individuals. The exercise programme followed a structured format, which included a warm-up, Otago exercise programme (OEP) [23], "workouts on-demand," and a cool-down. The same instructor, aged 66 and experienced in teaching older adults, conducted real-time demonstrations for both groups. The session was personalized for each individual. For instance, those with low muscle strength were given modifications by the instructor such as holding the exercise for a shorter duration or doing fewer repetitions. Feedback was provided to individual participants during and immediately after class on an as-needed basis.

### Intervention-tele-exercise group

For the TE group, the exercise programme was delivered via Zoom (Zoom Video Communications Inc. 2022) in the participants’ homes, allowing for real-time online communication with the participants. Participants without suitable devices were provided with all the necessary devices. Safety measures and technological support for the tele-exercise group Prior to the intervention, home visits were conducted to ensure a safe home environment, assist with setting up online communication applications, and optimize the positioning of the tablet, chair, and participant for effective visual interaction. The first lesson took place at the community centre to familiarize participants with exercise safety rules and the OEP, followed by online training sessions. Participants were reminded to turn on their tablets and Wi-Fi before each class so that we could control their tablet remotely to promptly address technical issues. Participants performed all OEP exercises, starting at the beginner level with support for standing and walking. Camera and microphone activation were required to enhance interaction, with regular progress monitoring by the research team. Participants were reminded to wear appropriate exercise gear and maintain hydration.

### Intervention-community-based group

For the CB group, the exercise classes were arranged at the community centres based on the residence of the recruited participants. The training programme was the same as that of TE group, except that it was conducted face-to-face at the community centre.

### Maintenance phase

After 3 months of training, workshops were arranged to remind the participants to maintain the exercise habits and maintain their current step count. If feasible, they were encouraged to gradually increase their daily step count by 3,500 steps. Exercise booklets from our exercise programme were distributed to all participants. The exercise videos were sent to them via WhatsApp, or presented in an easily accessible format. The links to the exercise booklet and exercise videos are available on the website of the Centre for Ageing and Healthcare Management Research [24]. They were reminded by phone at least twice per month about the exercise target and any falls incidence. Participants were required to report any instances of falls during the 12-month period.

### Outcome measures

Baseline assessments and post-intervention assessments at the three-month were conducted by trained and blinded assessors at the community centres. Participants received HK$100 gift vouchers at 3 month and 12 month for completing the training and assessments. In addition to the assessments, demographic information and background variables such as sex, age, and history of falls were collected. Cognitive function at baseline was evaluated using the Mini-Mental State Examination (MMSE) [25].

### Fall and functional outcomes

Fall efficacy was evaluated using the Chinese version of the Fall Efficacy Scale-International (FES-I) [26]. Participants were asked to proactively report any instances of falling and any adverse events during the tele-exercise sessions, follow-up assessments and the two monthly phone reminders. The severity of all fall incidences was classified based on the National Database of Nursing Quality Indicators® (NDNQI®) Injury Falls Measure [27]. Other outcomes included SARC-CalF, grip strength assessments for both the right and left hands, a 6-meter walk test, a timed up and go test (TUG), the Berg Balance Scale score (BBS), the functional reach test (FRT), appendicular skeletal muscle mass index (ASMI), percentage of body fat. All outcomes were measured at baseline, the 3^rd^ , 6^th^ , and 12^th^ month.

### Exercise experience and maintenance outcomes

The secondary outcomes evaluate older people’s involvement and experience of the exercise programmes including exercise adherence, dropout rate, Physical Activity Enjoyment Scale (PACES) [28], physical activity levels and step counts. The exercise adherence rate was defined as the percentage of attended classes out of the total 36 classes. The self-esteem of the participants was evaluated by the Rosenberg Self-Esteem Scale. Exercise maintenance was assessed using the Physical Activity Scale for the Elderly-Chinese (PASE-C) [29]. Physical activity levels and step counts were objectively measured using accelerometers (ActiGraph wGT3X-BT, Pensacola, FL, USA). Participants wore on their waist for seven consecutive days at baseline, and at the 3^rd^ , 6^th^ , and 12^th^ month. The ActiLife software was used to estimate their total steps and physical activity level based on the Freedson et al. [30] cut points.

### Data analysis

Baseline characteristics including age, adherence rate, and dropout rate, were analyzed using an independent t-test. The differences between categorical variables such as education level were analyzed by Chi-square test. An intention-to-treat analysis, following the guidelines for RCTs [31], was employed, including all originally randomized participants in the study. For the outcome measures, a Generalized Estimated Equation (GEE) Analysis with time and group as the main effects and baseline as a covariate. A pairwise comparison was conducted to assess the variances between the intervention groups and within-group variances across different time points. Post hoc Bonferroni correction was employed for the analysis of multiple outcomes. GEE can effectively manage missing data and provide an integrated approach to address missing values. Furthermore, effect sizes, expressed as Cohen’s d, were calculated. A significance level of 0.05 was set for all tests. The statistical analysis was carried out using SPSS, version 26.0 (IBM Co., Armonk, NY).

### Deviations from the registered trial protocol

Instead of the initially planned 92 participants, we recruited 94 participants as more dropout observed in TE, In terms of statistical analysis, we employed GEE instead of a two-way repeated measures ANOVA. This decision was made because GEE is more flexible with fewer assumptions about the data distribution and can handle missing data and unbalanced designs [32].

In the protocol, participants were initially instructed to achieve a daily step count of over 10,000 steps or increase their step count by 3,500 steps every three months until they reached the target of 10,000 steps per day. However, many participants expressed significant concerns about their busy schedules, with some citing caregiving responsibilities for family members as a barrier to meeting the required step count. Additionally, after practicing the prescribed exercise, they felt tired, which made it challenging for them to increase their step counts. As a result, they were more inclined to follow the OEP we recommended. To alleviate the pressure on participants, we revised our guidance for all groups. Participants were advised to perform the prescribed exercises at least three times per week and to maintain their current step count. If feasible, they were encouraged to gradually increase their daily step count by 3,500 steps.

The Physical Activity Enjoyment Scale (PACES) was utilized to evaluate the participants’ enjoyment of physical activity, which is a crucial factor in promoting sustained engagement in exercise programmes [33]. By incorporating PACES, the study aimed to obtain a more accurate and holistic understanding of enjoyment among older adults. The second objective of the protocol will also be analysed by qualitative analysis. Considering the large volume of data, it will be submitted as another manuscript.

## Results

### Flow of participants

During the screening period from October 2021 to June 2022, a total of 212 older adults were identified for preliminary screening. Out of these, 118 individuals were excluded based on the inclusion and exclusion criteria of the study. Eventually, 94 participants successfully met the inclusion criteria. The TE group comprised 48 participants, while the CB group consisted of 46 participants (Figure 1).

**Figure 1.**
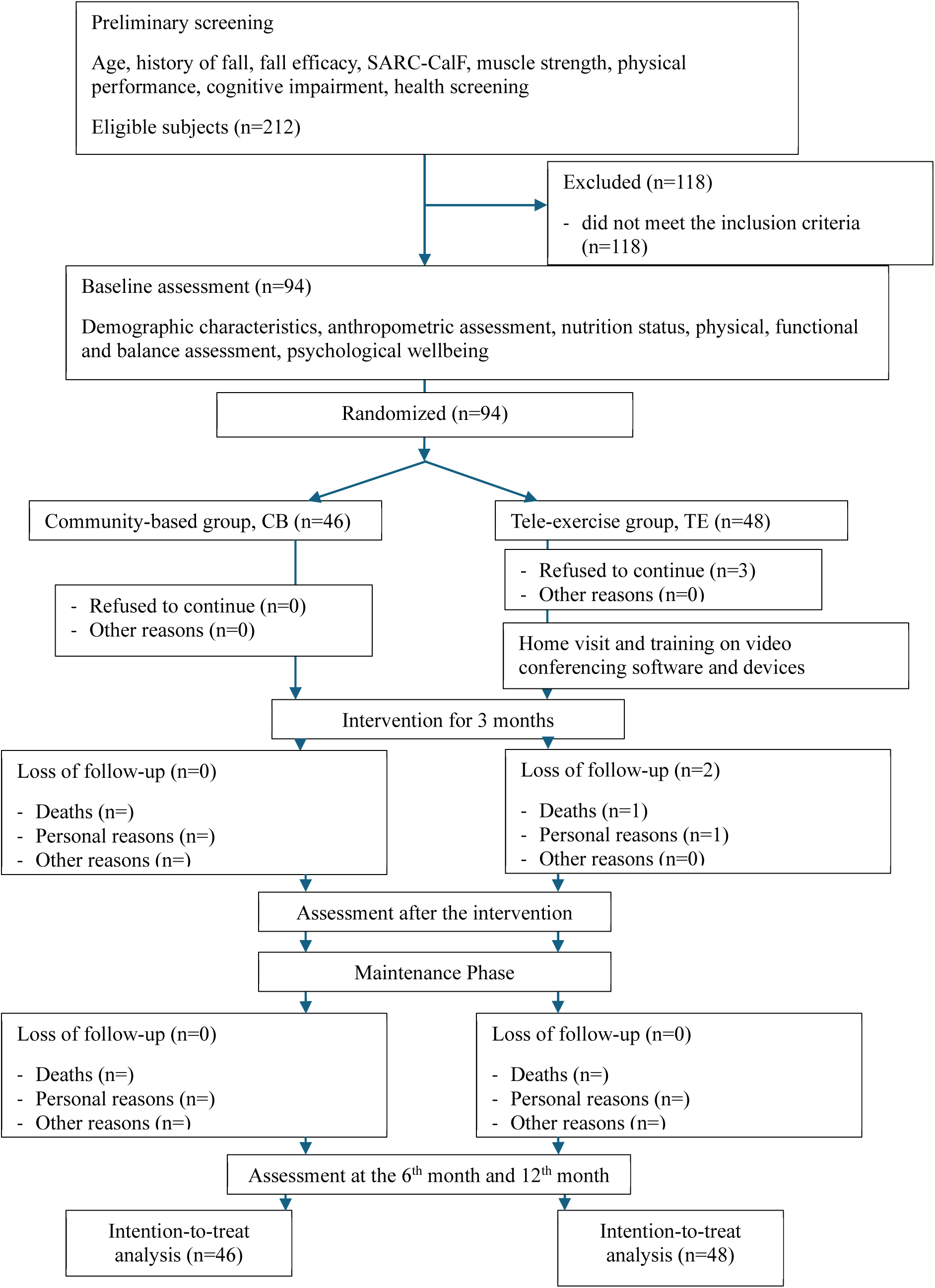
CONSORT flow chart

### Participant characteristics and education level

The baseline characteristics and education level of the participants did not show significant differences between the CB and TE groups, as summarized in Table 2.

**Table 2.** Demographic characteristics of the participants.

|  | Community-based (CB) | Tele-exercise (TE) |
| --- | --- | --- |
| Baseline Characteristics |  |  |
| n | 46 | 48 |
| Age (Range) | 77.3 ± 6.7 (65-93) | 75.0 ± 6.2 (65-91) |
| Proportion of female participants | 89.1% (41) | 87.5% (42) |
| Weight (kg) | 56.9 ± 12.1 | 58.0 ± 11.8 |
| Height (cm) | 150.3 ± 6.3 | 153.7 ± 8.1 |
| BMI (kg/m<sup>2</sup>) | 25.5 ± 5.2 | 24.6 ± 4.5 |
| Mini Nutritional Assessment (MNA) | 24.9 ± 2.0 | 25.0 ± 2.1 |
| Mini Mental State Examination (MMSE) | 27.59 ± 1.89 | 27.84 ± 1.78 |
| Education Level<sup>^</sup> |  |  |
| Junior primary or below | 47.8% (22) | 27.1% (13) |
| Senior primary | 21.7% (10) | 33.3% (16) |
| Junior Secondary | 21.7% (10) | 16.7% (8) |
| Senior Secondary | 8.7% (4) | 20.8% (10) |
| University or above | 0% (0) | 2.1% (1) |
| Technological accessibility<sup>^</sup> |  |  |
| Smartphone ownership | 71.7% (33) | 85.4%* (41) |
| Home Wi-Fi ownership | 67.4% (31) | 60.9% (28) |
| Previous experiences with tele-exercise | N/A | 14.6%(7) |

<sup>^</sup>Values represent percentage and number of participants.3 \**p*<0.05 compared with CB

### Technological accessibility and technological support for the TE participants

In terms of smartphone ownership, the TE group exhibited a higher percentage at 85.4%, while the CB group reported 71.7% (p<0.05). In terms of home Wi-Fi ownership, the CB group had a higher percentage at 67.4%, compared to the TE group at 60.9%, but this difference did not reach a significant level. Additionally, only 14.6% of participants in the TE group had previous experience with tele-exercise (Table 2).

Baseline home visits were conducted for all participants before the intervention and during the initial lessons, with an average of 3.7 visits per participant. During the initial lessons, student helpers were on standby at the homes of certain participants, especially those living alone. Subsequently, when participants faced technological issues, we typically resolved them before the following lesson, usually within 2 days. Additional visits addressed technological issues and provided support such as network connection problems, totaling 260 visits (including baseline visits) for the 45 participants in the TE group. On averaging, each participant received 5.8 visits (range: 0-12 visits), with a median of 5 visits.

### Falls incidence and fall risks

There was no significant difference between the groups in the incidence of falls. Prior to the intervention, the total number of falls was 71 in the CB and 80 in the TE over the past 12 months. The majority of falls were classified as minor incidents based on the guidelines from Garrard et al. [27]. During the intervention, the CB group reported no falls, whereas one participant in the TE group reported experiencing a fall outside of the lessons, which resulted in a moderate injury (Table 3). For within-group analysis, both the CB and TE groups exhibited a significant reduction in fall incidence at 12 months compared to baseline (p<0.05), with effect sizes of 1.03 for CB and 1.17 for TE, respectively. The FES-I scores improved significantly in both groups at 3^rd^ month (p<0.001), but only the CB group had a sustained effect at 6^th^ and 12^th^ month (Table 4-5).

**Table 3.** Falls incidence.

| Outcomes | Baseline<br>(Last 12<br>months<br>before<br>intervention) | 3<sup>rd</sup> month | 6<sup>th</sup> month | 12<sup>th</sup> month |
| --- | --- | --- | --- | --- |
| No. of participants in CB | 46 | 46 | 46 | 46 |
| No. of participants in TE | 48 | 43 | 43 | 43 |
| **Fall incidence** |  |  |  |  |
| **Total no. of falls** |  |  |  |  |
| CB (no. of participants) | 71 (46) | 0 (0) | 6 (6) | 9 (7) |
| TE (no. of participant) | 80 (48) | 1 (1) | 2 (2) | 11 (8) |
| **No. of minor injuries** |  |  |  |  |
| CB (no. of participant) | 61 (46) | 0 (0) | 3 (3) | 7 (6) |
| TE (no. of participant) | 74 (48) | 0 (0) | 1 (1) | 7 (5) |
| **No. of moderate injuries** |  |  |  |  |
| CB (no. of participant) | 8(6) | 0(0) | 1(1) | 0(0) |
| TE (no. of participant) | 4(4) | 1(1) | 1(1) | 3(3) |
| **No. of Serious injuries** |  |  |  |  |
| CB (no. of participant) | 2 (2) | 0 (0) | 2 (2) | 2 (2) |
| TE (no. of participant) | 2 (1) | 0 (0) | 0 (0) | 1 (1) |

^ The severity of all fall incidences was classified based on the NDNQI® Injury Falls Measure [27].

**Table 4.** Summary of Generalized Estimated Equation Analysis for primary and secondary outcomes.

| Outcomes | Mean (SD) [available data] |  |  |  | Between groups - P value<sup>a</sup> |  |  |
| --- | --- | --- | --- | --- | --- | --- | --- |
|  | Baseline | 3<sup>rd</sup> month | 6<sup>th</sup> month | 12<sup>th</sup> month | Interaction effect | Group effect | Time effect |
| **Fall and Functional Outcomes** |  |  |  |  |  |  |  |
| **Fall incidence** |  |  |  |  | 0.193 | 0.553 | <0.001 |
| CB | 1.5 (0.8) [46] |  |  | 0.3(0.7) [46] |  |  |  |
| TE | 1.6 (0.8) [48] |  |  | 0.3(0.7) [43] |  |  |  |
| **FES-I scores** |  |  |  |  | 0.876 | 0.852 | <0.001 |
| CB | 36.4(7) [46] | 33.4(8.1) [46] | 34.7(7.3) [46] | 35.1(7.6) [46] |  |  |  |
| TE | 35.4(7.7) [48] | 32.4(7.7) [43] | 34.2(7.2) [43] | 34.4(7.5) [43] |  |  |  |
| **SARC-CalF scores** |  |  |  |  | 0.426 | 0.542 | 0.008 |
| CB | 8(4.8) [46] | 6.9(4.5) [46] | 7.3(4.8) [46] | 7.2(4.9) [44] |  |  |  |
| TE | 8.8(4.6) [48] | 8.1(4.3) [42] | 8.0(4.6) [39] | 7.3(4.8) [41] |  |  |  |
| **6-meter Walk (m/s)** |  |  |  |  | 0.043 | 0.489 | <0.001 |
| CB | 0.7(0.2) [46] | 0.8(0.1) [46] | 0.8(0.1) [46] | 0.8(0.2) [44] |  |  |  |
| TE | 0.7(0.2) [48] | 0.8(0.2) [42] | 0.7(0.2) [40] | 0.7(0.3) [41] |  |  |  |
| **TUG (s)** |  |  |  |  | 0.010 | 0.057 | 0.002 |
| CB | 15.6(9.9) [46] | 13.9(11.7) [46] | 14.4(10.9) [46] | 14.9(13.1) [44] |  |  |  |
| TE | 13.1(5) [48] | 11.1(3.9) [43] | 9.8(3.6) [40] | 10.0(3.1) [40] |  |  |  |
| **BBS scores** |  |  |  |  | 0.820 | 0.964 | <0.001 |
| CB | 46.2(7.1) [46] | 49.7(9.5) [46] | 49.6(6.2) [45] | 49.3(8.2) [44] |  |  |  |
| TE | 46.9(8.1) [48] | 50.7(4.9) [43] | 50.7(6.2) [40] | 48.4(9.7) [41] |  |  |  |
| **FRT (cm)** |  |  |  |  | 0.129 | 0.989 | <0.001 |
| CB | 21.7(7.6) [46] | 24.4(6.4) [46] | 26.9(7.1) [46] | 26.9(9) [44] |  |  |  |
| TE | 26.7(6.6) [48] | 27(7.8) [43] | 28.4(7.9) [40] | 30.3(9.1) [41] |  |  |  |
| **Handgrip (kg)** |  |  |  |  | 0.417 | 0.963 | <0.001 |
| CB | 17(4.5) [46] | 17.9(4.8) [46] | 17.6(4.6) [46] | 18.6(4) [44] |  |  |  |
| TE | 19(6.1) [48] | 20.1(6.3) [42] | 19.8(6.3) [39] | 18.8(5.9) [41] |  |  |  |
| **ASMI kg/m<sup>2</sup>** |  |  |  |  | 0.609 | 0.521 | 0.756 |
| CB | 5.9(1.1) [46] | 5.9(1.1) [46] | 5.9(1.1) [46] | 6.1(1.2) [44] |  |  |  |
| TE | 6.2(1.0) [47] | 6.4(1.8) [41] | 6.2(1.0) [38] | 6.2(0.9) [40] |  |  |  |
| **% Body Fat** |  |  |  |  | 0.299 | 0.106 | 0.001 |
| CB | 36.0(6.6) [46] | 36.8(9.5) [46] | 35.4(7.1) [46] | 34.9(6.4) [44] |  |  |  |
| TE | 34.0(8.3) [47] | 33.3(8) [41] | 33.0(7.4) [38] | 33.2(7.9) [40] |  |  |  |
| **Exercise Experience and Maintenance Outcomes** |  |  |  |  |  |  |  |
| **Rosenberg Self-Esteem Scale** |  |  |  |  | 0.289 | 0.833 | 0.092 |
| CB | 21.6(3.3) [46] | 21.3(2.6) [46] | 20.6(3.2) [46] | 21.8(3.3) [46] |  |  |  |
| TE | 21.9(3.3) [48] | 21.9(3.7) [41] | 21.2(3.6) [43] | 20.9(4.3) [40] |  |  |  |
| **PASE-C (min)** |  |  |  |  | 0.001 | <0.001 | <0.001 |
| CB | 122.0(65.3) [46] | 157.6(64.7) [46] | 147.5(62.1) [46] | 164.0(66.7) [46] |  |  |  |
| TE | 115.8(63.5) [48] | 154.7(62.1) [43] | 151.3(62.1) [43] | 178.0(67.6) [42] |  |  |  |
| **Total steps (steps/day)** |  |  |  |  | 0.639 | 0.375 | 0.215 |
| CB | 5053.2(3445.3) [46] | 5077.0(3160.1) [46] | 4982.2(3082.4) [46] | 4901.5(3081) [42] |  |  |  |
| TE | 6437.5(3121.7) [48] | 6355.6(3383.7) [41] | 6883.3(3570.8) [39] | 5944.1(2577.3) [38] |  |  |  |
| **Sedentary (min/day)** |  |  |  |  | 0.337 | 0.082 | 0.078 |
| CB | 1240.2(199.2)<br>[46] | 1238.1(71.1)<br>[46] | 1228.2(177.5)<br>[46] | 1251.3(88.9)<br>[42] |  |  |  |
| TE | 1212.5(99.4)<br>[48] | 1175.9(215.1)<br>[41] | 1197.8(110.8)<br>[39] | 1240.1(53.9)<br>[38] |  |  |  |
| **Light activity (min/day)** |  |  |  |  | <0.001 | 0.270 | <0.001 |
| CB | 152.6(56.6)<br>[46] | 165.6(57.1)<br>[46] | 167.8(75.5)<br>[46] | 186.7(86.3)<br>[42] |  |  |  |
| TE | 161.9(58.4)<br>[48] | 127.5(79)<br>[41] | 245.7(106.6)<br>[39] | 229.6(321.3)<br>[38] |  |  |  |
| **MVPA (min/day)** |  |  |  |  | 0.361 | 0.637 | 0.002 |
| CB | 16.7(17.5)<br>[46] | 16.6(19.3)<br>[46] | 28.2(61.8)<br>[46] | 12.9(15.6)<br>[42] |  |  |  |
| TE | 25.2(25.5)<br>[48] | 25.8(29) [41] | 24(22.7) [39] | 22.1(16.8)<br>[38] |  |  |  |

<sup>a</sup> Generalized estimated equation model, with baseline measurement as the covariate, was used to analyze the data.
FES-I: Fall Efficacy Scale; TUG: Timed Up and Go Test; BBS: Berg Balance Scale score; FRT: Functional Reach Test;
Appendicular Skeletal Muscle Mass Index: ASMI; PASE-C: Chinese version of the Physical Activity Scale for the Elderly;

### Physical function and muscle mass

The TE group showed a significantly higher ASMI at the 6th month compared to the CB group (p<0.05, effect size: 0.00). No significant between-group differences were found for other physical function measures across the 3^rd^, 6^th^, and 12^th^ months (Table 5).

**Table 5.** Summary of Post Hoc Analysis of Generalized Estimated Equation Analysis for primary and secondary outcomes (Bonferroni correction)

|  | Within-group differences |  |  |  |  |  | Between group differences |  |  |  |  |  |
| --- | --- | --- | --- | --- | --- | --- | --- | --- | --- | --- | --- | --- |
|  | Baseline vs 3<sup>rd</sup> month |  | Baseline vs 6<sup>th</sup> month |  | Baseline vs 12<sup>th</sup> month |  | 3<sup>rd</sup> month |  | 6<sup>th</sup> month |  | 12<sup>th</sup> month |  |
|  | Mean<br>adjusted<br>change<br>(95% CI) | Effect size<sup>b</sup> | Mean<br>adjusted<br>change<br>(95% CI) | Effect size<sup>b</sup> | Mean<br>adjusted<br>change<br>(95% CI) | Effect size<sup>b</sup> | Mean<br>adjusted<br>change<br>(95% CI) | Effect size<sup>b</sup> | Mean<br>adjusted<br>change<br>(95% CI) | Effect size<sup>b</sup> | Mean<br>adjusted<br>change<br>(95% CI) | Effect size<sup>b</sup> |
| **Fall and Functional Outcomes** |  |  |  |  |  |  |  |  |  |  |  |  |
| **Fall incidence** |  |  |  |  |  |  |  |  |  |  |  |  |
| CB |  |  |  |  | -1.24(-1.6 to -0.9)* | 1.03 |  |  |  |  | -0.16 (-0.6 to 0.3) | 0.11 |
| TE |  |  |  |  | -1.40(-1.7 to -1.1)* | 1.17 |  |  |  |  |  |  |
| **FES-I scores** |  |  |  |  |  |  |  |  |  |  |  |  |
| CB | -3.04(-4.5 to -1.5)* | 0.65 | -1.70(-2.5 to -0.8)* | 0.64 | -1.28(-2.3 to -0.2)* | 0.40 | -0.02 (-2.9 to 2.9) | 0.00 | 0.42 (-3.1 to 3.9) | 0.04 | -0.27 (-3.9 to 3.4) | 0.02 |
| TE | -2.90(-5.4 to -0.4)* | 0.38 | -1.11(-4.7 to 2.5) | 0.10 | -0.86(-4.5 to 2.8) | 0.08 |  |  |  |  |  |  |
| **SARC-CalF scores** |  |  |  |  |  |  |  |  |  |  |  |  |
| CB | -1.13(-2.6 to 0.3) | 0.25 | -0.70(-2.4 to 1) | 0.13 | -0.81(-2.5 to 0.9) | 0.16 | 0.66 (-0.8 to 2.1) | 0.21 | 0.43 (-1.7 to 2.6) | 0.29 | 0.23 (-2.4 to 2.9) | 0.00 |
| TE | -0.63(-1.4 to 0.1) | 0.28 | -0.42(-2.1 to 1.2) | 0.08 | -1.20(-3.5 to 1.1) | 0.17 |  |  |  |  |  |  |
| **6-meter Walk (m/s)** |  |  |  |  |  |  |  |  |  |  |  |  |
| CB | 0.06(0.0 to 0.1)* | 0.40 | 0.08(0.0 to 0.1)* | 0.34 | 0.06(0.0 to 0.1) | 0.26 | 0.02 (-0.1 to 0.1) | 0.04 | -0.05 (-0.1 to 0.1) | 0.18 | -0.02 (-0.1 to 0.1) | 0.07 |
| TE | 0.08(0.0 to 0.1)* | 0.52 | 0.03(0.0 to 0.1) | 0.16 | 0.05(0.0 to 0.1) | 0.18 |  |  |  |  |  |  |
| **TUG (s)** |  |  |  |  |  |  |  |  |  |  |  |  |
| CB | -1.71(-4.2 to 0.8) | 0.22 | -1.17(-4.0 to 1.6) | 0.14 | -0.61(-3.7 to 2.5) | 0.06 | -0.35 (-2.8 to 2.1) | 0.08 | -1.30 (-4.2 to 1.6) | 0.17 | -2.68 (-5.9 to 0.5) | 0.28 |
| TE | -1.95(-3.4 to 0.5)* | 0.42 | -2.37(-4.0 to 0.8) | 0.47 | -3.18(-5.2 to 1.2)* | 0.51 |  |  |  |  |  |  |
| **BBS scores** |  |  |  |  |  |  |  |  |  |  |  |  |
| CB | 3.54(-0.4 to 7.5) | 0.29 | 3.12(0.5 to 5.7)* | 0.39 | 2.99(-0.4 to 6.4) | 0.28 | 0.33 (-3.8 to 4.5) | 0.00 | 0.52 (-2.9 to 3.8) | 0.03 | -1.22 (-6.9 to 4.3) | 0.09 |
| TE | 3.62(1.1 to 6.1)* | 0.47 | 3.4(0.4 to 6.4)* | 0.36 | 1.52(-3.4 to 6.4) | 0.10 |  |  |  |  |  |  |
| **FRT (cm)** |  |  |  |  |  |  |  |  |  |  |  |  |
| CB | 2.74(-0.3 to 5.8) | 0.29 | 5.22(1.5 to 8.9)* | 0.46 | 5.3(1.5 to 9.1)* | 0.44 | 0.54 (-3.5 to 4.6) | 0.07 | 1.69 (-2.9 to 6.2) | 0.14 | 0.32 (-5.6 to 6.9) | 0.01 |
| TE | 0.33(-3.1 to 3.8) | 0.03 | 1.67(-1.6 to 5) | 0.16 | 3.76(-1 to 8.6) | 0.25 |  |  |  |  |  |  |
| **Handgrip (kg)** |  |  |  |  |  |  |  |  |  |  |  |  |
| CB | 0.95(-0.1 to 2) | 0.28 | 0.67(-0.9 to 2.2) | 0.14 | 1.46(-0.6 to 3.5) | 0.23 | 0.28 (-1.1 to 1.7) | 0.01 | 0.29 (-2.1 to 2.7) | 0.01 | 0.97 (-1.7 to 3.7) | 0.15 |
| TE | 0.91(0.1 to 1.7)* | 0.36 | 0.63(-1.3 to 2.6) | 0.11 | -0.16(-2.3 to 2.0) | 0.02 |  |  |  |  |  |  |
| **ASMI (kg/m<sup>2</sup>)** |  |  |  |  |  |  |  |  |  |  |  |  |
| CB | -0.04(-0.2 to 0.1) | 0.08 | -0.01(-0.2 to 0.2) | 0.02 | 0.11(-0.3 to 0.6) | 0.08 | 0.23 (-0.4 to 0.8) | 0.10 | 0.03 (-0.1 to 0.2)^ | 0.00 | 0.09 (-0.3 to 0.5) | 0.09 |
| TE | 0.17(-0.5 to 0.8) | 0.09 | 0.00(-0.1 to 0.1) | 0.00 | 0.00(-0.1 to 0.1) | 0.00 |  |  |  |  |  |  |
| **% Body Fat** |  |  |  |  |  |  |  |  |  |  |  |  |
| CB | 0.83(-1.8 to 3.5) | 0.10 | -0.57(-1.7 to 0.5) | 0.17 | -1.00(-2.2 to 0.2) | 0.27 | -1.86 (-4.9 to 1.2) | 0.18 | -0.83 (-3.0 to 1.4) | 0.12 | -0.47 (-2.6 to 1.7) | 0.06 |
| TE | -0.91(-2.4 to 0.6) | 0.20 | -1.29(-3.1 to 0.5) | 0.23 | -1.35(-3.0 to 0.3) | 0.26 |  |  |  |  |  |  |
| **Exercise Experience and Maintenance Outcomes** |  |  |  |  |  |  |  |  |  |  |  |  |
| **Rosenberg Self-Esteem Scale** |  |  |  |  |  |  |  |  |  |  |  |  |
| CB | -0.24(-1.9 to 1.4) | 0.05 | -0.96(-2.7 to 0.8) | 0.17 | 0.20(-1.7 to 2.1) | 0.03 | 0.27 (-2.6 to 1.7) | 0.04 | 0.33 (-1.8 to 2.5) | 0.06 | -1.11 (-3.6 to 1.4) | 0.14 |
| TE | -0.12(-2.3 to 2.1) | 0.02 | -0.78(-2.6 to 1.1) | 0.14 | -1.08(-3.2 to 1.1) | 0.16 |  |  |  |  |  |  |
| **PASE-C (min)** |  |  |  |  |  |  |  |  |  |  |  |  |
| CB | 35.61(32.7 to 38.5)* | 3.94 | 25.5(18.4 to 32.6)* | 1.15 | 41.93(34.8 to 49.1)* | 1.89 | -2.27 (-5.22 to 0.68) | 0.27 | 8.94 (-0.72 to 18.6) | 0.32 | 21.48 (2.5 to 40.5)^ | 0.37 |
| TE | 38.02(37.8 to 38.3)* | 46.62 | 34.58(27.9 to 41.3)* | 1.66 | 63.56(46.0 to 81.1)* | 1.16 |  |  |  |  |  |  |
| **Total steps (steps/day)** |  |  |  |  |  |  |  |  |  |  |  |  |
| CB | 23.83(-1121.7 to 1169.4) | 0.01 | -71.02(-1024.3 to 882.3) | 0.02 | -244.88(-1401.0 to 911.2) | 0.07 | 199.25 (-1226.4 to 1624.9) | 0.06 | 593.72 (-833.8 to 2021.2) | 0.15 | 45.74 (-1461.4 to 1552.9) | 0.00 |
| TE | -59.13(-960.0 to 841.7) | 0.02 | 240.5(-860.2 to 1341.2) | 0.07 | -572.82(-1715.1 to 569.5) | 0.16 |  |  |  |  |  |  |
| **Sedentary (min/day)** |  |  |  |  |  |  |  |  |  |  |  |  |
| CB | -2.1(-102.1 to 97.9) | 0.01 | -11.98(-128.8 to 104.8) | 0.03 | 3.27(-74.7 to 81.3) | 0.01 | -53.46 (-167.86 to 60.94) | 0.15 | -26.2 (-127.2 to 74.79) | 0.09 | 6.01 (-45.18 to 57.21) | 0.03 |
| TE | -37.46(-146.6 to 71.7) | 0.11 | -20.09(-86.1 to 45.9) | 0.10 | 27.37(-18.5 to 73.2) | 0.19 |  |  |  |  |  |  |
| **Light activity (min/day)** |  |  |  |  |  |  |  |  |  |  |  |  |
| CB | 13.05(-15.3 to 41.4) | 0.15 | 15.18(-23.3 to 53.6) | 0.13 | 31.71(-11 to 74.4) | 0.24 | -44.45 (-89.95 to 1.04) | 0.31 | 66.57 (9.06 to 124.08)^ | 0.33 | 38.14 (-127.03 to 203.31) | 0.08 |
| TE | -35.03(-78.3 to 8.2) | 0.26 | 78.12(34 to 122.3)* | 0.57 | 66.21(-94.4 to 226.8) | 0.13 |  |  |  |  |  |  |
| **MVPA (min/day)** |  |  |  |  |  |  |  |  |  |  |  |  |
| CB | -0.16(-5.5 to 5.2) | 0.01 | 11.44(-13.5 to 36.3) | 0.15 | -4.23(-8.7 to 0.2)* | 0.31 | 2.15 (-12.09 to 16.39) | 0.06 | -11.09 (-39.58 to 17.4) | 0.12 | 2.14 (-5.59 to 9.87) | 0.10 |
| TE | 1.7(-11.3 to 14.7) | 0.04 | -0.05(-8 to 8.1) | 0.00 | -2.39(-8 to 3.2) | 0.14 |  |  |  |  |  |  |

.FES-I: Fall Efficacy Scale; TUG: Timed Up and Go Test; BBS: Berg Balance Scale score; FRT: Functional Reach Test; PASE-C: Chinese version of the Physical Activity Scale for the Elderly;
<sup>b</sup>Effect size: Mean difference/SD
\* $p < 0.05$ for within group difference compared with baseline.
<sup>^</sup> $p < 0.05$ for between group difference.

Within-group analyses revealed significant improvements in physical function and physical activity across multiple time points for both groups. In the TE group, significant improvements at the 3^rd^ month were observed in the 6-meter walk test (p<0.05, effect size: 0.52), TUG scores (p<0.05, effect size: 0.42), BBS scores (p<0.05, effect size: 0.47), and handgrip strength (p<0.05, effect size: 0.36). The TE group also showed significant improvements in TUG at the 12th month (p<0.05, effect size: 0.51) and BBS at the 6th month (p<0.05, effect size: 0.36). For the CB group, significant improvements were noted in the 6-meter walk test at the 3rd (p<0.05, effect size: 0.40) and 6th months (p<0.05, effect size: 0.34), BBS at the 6th month (p<0.05, effect size: 0.39), and FRT at the 6th (p<0.05, effect size: 0.46) and 12th months (p<0.05, effect size: 0.44).

### Exercise experience and exercise maintenance

The CB group experienced a significantly higher adherence rate compared with the TB group (CB: 90% vs TE: 85%, p=0.01). In terms of dropout rates, the CB group had a 0% dropout rate, whereas the TE group had a dropout rate of 10.4% (p<0.01).

The PACES scores were comparable between the two groups, with the CB group scoring 108.2 (range: 80–126) and the TE group scoring 108.6 (range: 95–121) out of a maximum of 126 (p > 0.05). Additionally, there was no significant difference between the groups in the Rosenberg Self-Esteem Scale.

The TE group showed significantly higher PASE-C scores than the CB group at 12 months (p<0.05, ES=0.37). Both groups exhibited significant within-group PASE-C increases from baseline at 3 months (CB: ES=3.94; TE: ES=46.62), 6 months (CB: ES=1.15; TE: ES=1.66), and 12 months (CB: ES=1.89; TE: ES=1.16; all p<0.05). TE had significantly higher light activity than CB at 6 months (p<0.05, ES=0.33), with TE also showing a significant within-group increase in light activity at 6 months from baseline (ES=0.57, p<0.05). CB displayed significantly lower moderate-to-vigorous physical activity (MVPA) at 12 months compared to baseline (ES=0.31, p<0.05). No significant between-group or within-group differences were found for total step count across the study period.

### Adverse events reporting

One participant in the TE group reported general tiredness and pain during screening phase, but her doctor found no issues after thorough examinations and tests. Living alone and rarely engaging in social activities, she insisted on participating in our programme to maintain her physical functioning.

During the class, she reported tiredness, knee and lower back pain at the beginning. The instructor advised her to rest when needed and practice at an easier level. With the frequent reminders to lower the intensity, her problems subsided. She completed the programme with an adherence rate of 72.2%.

## Discussion

### Key findings

To our knowledge, this study is the first rigorously designed RCT with sufficient statistical power to investigate the effectiveness of the synchronous group-based tele-exercise compared with the community-based group in older adults at risk of falls. The TE group demonstrated a slight advantage in exercise maintenance than the CB group. The TE group reported significantly higher PASE-C scores than the CB group (p<0.05, ES=0.37) at 12 months, with increases of 63.56 minutes for TE and 41.93 minutes for CB, reflecting greater perceived engagement in diverse activities including household tasks, leisure, and OEP exercises. At 6 months, the TE group exhibited significantly higher light physical activity than the CB group (p<0.05, ES=0.33), with a notable within-group increase from baseline (ES=0.57, p<0.05). Moreover, moderate-to-vigorous physical activity (MVPA) was better maintained in the TE group throughout the 12-month period. This sustained exercise maintenance may be attributed to participants’ adaptation to home-based exercise during the initial three-month intervention, facilitating independent continuation over the subsequent nine months.

The discrepancy between self-reported PASE-C gains and relative stagnant Actigraph metrics underscores the strengths and limitations of each approach. PASE-C effectively captured diverse activity profiles, including non-ambulatory and low-intensity tasks, which are critical for older adults [29]. The accelerometer used in the study may have underestimated our participants’ step counts due to their average low walking speeds (0.7–0.8 m/s) as the Actigraph sensor has reduced accuracy at low walking speeds (<0.9 m/s) [34]. Protocol deviations influenced outcomes, with participants citing barriers to increasing steps. Revised guidance to maintain current step counts and prioritize OEP exercises three times weekly likely sustained PASE-C improvements by focusing on structured, functional exercises. This shift supported functional outcomes such as reduced fall incidence, improved FES-I, FRT, and TUG scores but limited step count increases, as daily ambulation remained largely unchanged. Objective measures like Actigraph step counts require devices validated for slower walking speeds [34, 35].

The study found that the tele-exercise (TE) group had a significantly lower adherence rate (80% vs. 90%) and a higher dropout rate (10.4% vs. 0%) compared to the community-based (CB) group among older adults at risk of falls. Excluding three TE participants who attended no sessions due to family objections or concerns about unsafe home environments, adherence improved to 85%, surpassing the approximately 70% reported in community-based exercise programmes [36]. Of the two remaining dropouts, one passed away, and the other cited repetitive, unengaging course content, highlighting the need for varied and stimulating exercise programmes to sustain participation. These findings suggest skepticism toward tele-exercise among some older adults and their families, likely due to technological barriers or safety concerns. Additionally, the absence of in-person interaction in TE may reduce social motivation and accountability, key drivers of exercise maintenance in face-to-face settings [37]. Despite these challenges, the TE group achieved comparable improvements in physical function, indicating effectiveness for those who persisted. Success factors included professional supervision, individualized technology orientation, and prompt technical support (averaging 5.8 home visits per participant, 16.1% of 36 sessions), which ensured safety, effective problem-solving, and no serious adverse events, demonstrating the efficacy and safety of synchronous delivery of the modified Otago Exercise Programme [38, 39].

The study findings indicate that both the CB and TE interventions were comparable and effective in decreasing fall incidence. These results regarding the effectiveness of the modified OEP are consistent with the findings of systematic review conducted by Yang et al. [40] and Wu et al. [41]. The positive outcomes can be attributed to the fact that our intervention adhered to the recommended effective programme requirements identified by Wu et al. [41], with an average duration of 45 minutes, three times per week for 12 weeks. The consistent delivery of these components likely contributed to the observed reductions in fall incidence, highlighting the robustness of both CB and TE approaches in enhancing physical function and safety.

The sustained effects of our group-based interventions on fear of falling and functional outcomes varied between the CB and TE groups. The CB group showed significant within-group reductions from baseline at 3 months (-3.04 points, effect size = 0.65), 6 months (-1.70 points, effect size = 0.64), and 12 months (-1.28 points, effect size = 0.40; p<0.05). Conversely, the TE group exhibited a significant reduction only at 3 months (-2.9 points, effect size = 0.38; p<0.05), with non-significant changes thereafter. Using baseline FES-I data (CB: 36.4±7; TE: 36.5±7.7; pooled SD≈7.2), we estimated an MCID of 3.6 points via the 0.5 SD method [42]. CB’s 3-point reduction (8.35% improvement, ES=0.65) nears this threshold, while TE’s 2.9-point reduction (8.19% improvement, ES=0.38) falls short [43]. Although no anchor-based MCID exists for FES-I in this population [44], CB’s changes, with medium-to-large ES, suggest clinically meaningful improvements, especially given high baseline scores (mean=35.4–36.5). CB’s consistent FES-I and FRT improvements, plus higher adherence, highlight the value of face-to-face interactions.

### Bridging technological and education gaps in tele-exercise

Most tele-exercise studies primarily recruited individuals with existing internet access at home, favoring those who were technologically competent [11–13, 37]. A strength of our study was that technological competence and device ownership were not inclusion criteria. We provided participants with the necessary devices, welcoming individuals with various technological backgrounds. Our sample consisted of less educated individuals, with over 50% completed primary education or below, and only one TE participant completing university. Our findings indicated that older adults, regardless of education and technological proficiency, could enjoy tele-exercise when accompanied by well-designed programmes, technical support, and timely feedback. In alignment with Lee et al. [45], even individuals classified as the "oldest old," aged over 80, navigated a process of technological adaptation. They faced challenges, explored new avenues, and persistently adjusted to the technological facets of tele-exercise, highlighting the crucial role of technical support in their adaptation process. Furthermore, the utilization of commonly available telecommunications software in the trial, instead of custom-built systems [13], enhanced the generalizability of the findings [46].

### Limitations

One challenge of the study was the limited ability to capture full-body images due to spatial constraints in participants’ homes. Census and Statistics Department [47] indicate that the median floor areas for domestic households and individuals in Hong Kong are 40 and 16 m^2^, respectively. This spatial constraint could impact the ability to capture a full view of participants while they were standing during tele-exercise sessions. To overcome this issue, we made efforts during home visits to find the optimal placement and angle for tablets with built-in cameras to maximize visibility during the exercise sessions. To ensure understanding of the exercise programme and promote exercise safety, all participants were required to attend the first class at the community centre. This allowed for in-person instructions and reminders before transitioning to tele-exercise sessions [18]. It is important to note that the success of this programme may not be transferable to other exercises that involve more dynamic movements such as standing, sitting, and lying down, within a single lesson. From a technological standpoint, it would be beneficial for the technology company to explore the possibility of incorporating tablets with wide-angle front cameras or establishing a seamless connection between a tablet and an external camera without any delays. These enhancements would help improve visibility during tele-exercise sessions.

The second limitation concerns the impact of multiple testing. Assessing multiple outcomes, including the FES-I, 6-meter walk test, TUG, BBS, FRT, ASMI, percentage of body fat, exercise adherence, dropout rate, and PACES, heightens the risk of Type I errors. This increases the likelihood of inflated false-positive rates, posing a methodological concern. To address this, we adopted a modified intention-to-treat approach, incorporating all available data and applying Bonferroni adjustments to control for multiple comparisons.

## Conclusion

This rigorously designed RCT, the first with sufficient statistical power to compare synchronous group-based tele-exercise (TE) with community-based (CB) interventions, demonstrates their comparable efficacy in enhancing physical functioning (e.g., fall efficacy, balance, mobility) and reducing fall incidence among older adults at risk of falls (aged 65–91), including those with varied education and limited tele-exercise experience. TE showed a slight advantage in sustaining long-term exercise maintenance, likely due to participants’ adaptation to home-based training, but its lower adherence due to higher dropout reflects skepticism towards tele-exercise, underscoring the need for robust technological support and safety measures. Clinically, synchronous tele-exercise can be integrated into fall prevention programmes, particularly when in-person access is limited, with support for devices, Wi-Fi, and tailored supervision to ensure safety and engagement. Policymakers should fund such initiatives to improve older adults’ health, whilst addressing barriers like skepticism through education and technologies such as wide-angle cameras, and inclusive interventions.

## Data Availability

All data produced in the present study are available upon reasonable request to the authors

## List of abbreviations

ASMI: Appendicular Skeletal Muscle Mass Index
BBS: Berg balance scale
CB: Face-to-face community-based exercise training
FES-I: Falls efficacy scale international
FRT: Functional Reach Test
GEE: Generalized Estimating Equations
MMSE: Mini-Mental State Examination
OEP: Otago exercise programme
RCT: Randomized controlled trial
PACES: Physical Activity Enjoyment Scale
PASE-C: Physical Activity Scale for the Elderly
TE: Synchronous group-based tele-exercise training
TUG: Timed Up and Go

## Declarations

### Ethics approval and consent to participate

The study conforms to the Declaration of Helsinki. The study involves human participants and was approved by the Ethics Committee of the College of Professional and Continuing Education, The Hong Kong Polytechnic University (RC/ETH/H/0029). All participants provided informed consent to participate in the study prior to their involvement.

### Consent for publication

Obtained.

### Availability of data and materials

The datasets used and analysed during the current study are available from the corresponding author on reasonable request.

### Author’s contribution

KOWC, PPY, BYFF, VTSL, FSFN, WCPF and TKCN conceived the study. KOWC and JLCC wrote the main manuscript text. ISC collected the data. KOWC and ISC performed statistical analysis. All authors reviewed the manuscript and approved the final version of the manuscript.

## Acknowledgements

The authors are grateful to the Centre for Ageing and Healthcare Management Research (CAHMR), College of Professional and Continuing Education, The Hong Kong Polytechnic University for coordination with the elderly services providers i.e. Sik Sik Yuen. Thank Sik Sik Yuen for recruiting older people and all the participants for joining this study. The authors would like to express their gratitude to Mr. Au Wing Ho for his exceptional exercise teaching provided to all the participants. Additionally, the authors extend their heartfelt thanks to the student helpers who offered technical support to the participants.

